# Repeat SARS-CoV-2 Testing Models for Residential College Populations

**DOI:** 10.1101/2020.07.09.20149351

**Authors:** Joseph T. Chang, Forrest W. Crawford, Edward H. Kaplan

## Abstract

Residential colleges are considering re-opening under uncertain futures regarding the COVID-19 pandemic. We consider repeat SARS-CoV-2 testing models for the purpose of containing outbreaks in the residential campus community. The goal of repeat testing is to detect and isolate new infections rapidly to block transmission that would otherwise occur both on and off campus. The models allow for specification of aspects including scheduled on-campus resident screening at a given frequency, test sensitivity that can depend on the time since infection, imported infections from off campus throughout the school term, and a lag from testing until student isolation due to laboratory turnaround and student relocation delay. For early- (late-) transmission of SARS-CoV-2 by age of infection, we find that weekly screening cannot reliably contain outbreaks with reproductive numbers above 1.4 (1.6) if more than one imported exposure per 10,000 students occurs daily. Screening every three days can contain outbreaks providing the reproductive number remains below 1.75 (2.3) if transmission happens earlier (later) with time from infection, but at the cost of increased false positive rates requiring more isolation quarters for students testing positive. Testing frequently while minimizing the delay from testing until isolation for those found positive are the most controllable levers for preventing large residential college outbreaks. A web app that implements model calculations is available to facilitate exploration and consideration of a variety of scenarios.

## 1 Introduction

Universities and colleges around the world, along with other businesses and institutions, have spent the past several months on lockdown on account of the COVID-19 pandemic. Students were sent home, classes and faculty meetings went on-line, and university buildings have remained eerily empty. With stay-at-home restrictions being relaxed if not rescinded, residential universities and colleges are planning to re-open, and perhaps the most prominent decision to be made is whether or not to invite students to return in person. Different schools are approaching this issue in different ways. Some colleges feel they have no choice but to allow students to return (Daniels 2020, Jenkins 2020), while others have opted for students to remain off-campus with instruction offered remotely via the internet (Castle 2020, Laframboise 2020). At the heart of this decision is the anticipated ability of colleges and universities to keep campuses free from student-driven SARS-CoV-2 outbreaks. While the details of infection control and social distancing represent important public health components on campus, increasingly institutions are considering whether testing students for SARS-CoV-2 offers additional protection against the spread of infection (Anderson and Svriuga 2020, Diep and Zahneis 2020, UCSD 2020).

Viral testing for SARS-CoV-2 infections is different than standard diagnostic testing. Usually when a patient is tested for the presence of some medical condition, it leads to a specific set of instructions for the benefit of the patient screened: a change in diet, the prescription of drugs, or a course of more intensive medical treatment such as radiation, chemotherapy, or surgery. With coronavirus, however, the purpose of testing is not to gain access to some treatment. Rather, those who test positive are instructed to isolate to prevent transmitting infection to others. The purpose of repeatedly screening students for SARS-CoV-2 is thus not for the screened patient’s individual health, but for the benefit of those who would have been in contact with an infectious person had the detection of an infection not occurred.

It is well-documented that contagious infections such as influenza, mumps, and sexually transmitted diseases spread readily in the residential college context (Bauer-Wolf 2019, Costill and Muoio 2015, Mangan 2019), and there is no reason to expect that SARS-CoV-2 would not be transmitted as well. However, students themselves are not at great medical risk from COVID-19 complications resulting from infection with SARS-CoV-2. Indeed, many if not most students would experience mild to no symptoms of infection at all. However, absent testing, all such infected students would unknowingly pose risks to anyone with whom they are in contact, whether on campus or off. For residential colleges that are themselves isolated geographically, vulnerable workers and faculty (and some students with underlying health conditions to be sure) are the main beneficiaries of repeat student testing. For urban campuses centrally located within surrounding communities that contain many more vulnerable persons, the main beneficiary of screening students is that community itself. In this sense, beyond protecting the health of vulnerable workers, faculty and students, the main goal of repeatedly screening students on campus is to prevent them from unknowingly igniting transmission chains in the surrounding community.

The way screening programs work to impact the transmission of infection in this context has not been well studied or analyzed. This paper presents a first attempt to do so. We begin with a simple characterization of the early transmission dynamics associated with nascent outbreaks of SARS-CoV-2, and in Section 3 show how test frequency, sensitivity and reporting delay influence transmission via isolating those testing positive when test results are obtained. In Section 4 we incorporate this interruption of transmission directly into a dynamic model of an internally generated SARS-CoV-2 outbreak on campus, and we expand the model to include exposures to imported infections from off-campus due to students traveling, wandering about town to restaurants, bars or clubs, or due to visitors. We present the key performance measures by which to assess repeat screening focusing on the cumulative incidence of infection, the number of infections detected, and the number of students placed in isolation for given outbreak and screening scenarios (different reproductive numbers governing on-campus transmission, different imported exposure rates, different screening frequencies, different test sensitivity, specificity, and delay from testing until isolation for those testing positive). We consider numerous scenarios in Section 5 with a focus on which outbreaks can and cannot be brought under control. We discuss implications of our analysis in Section 6.

## 2 Age-of-Infection Dependent Transmission

The model employed to analyze repeat screening follows Kaplan (2020a) in presuming that at the beginning of an outbreak, a newly-infected index surrounded by otherwise uninfected students transmits infections according to a time-varying Poisson process with intensity *λ*(*a*), where *a* denotes the time from infection of the index (the *age* of infection). The reproductive number denoting the expected number of infections the index will transmit over all time then

Equals

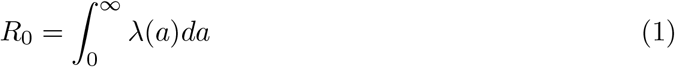

and as is well-known, an epidemic cannot be self-sustaining unless *R*_0_ *>* 1. The transmission intensity *λ*(*a*) can be represented as

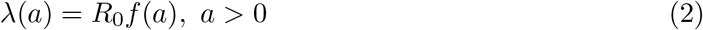

where

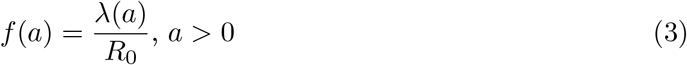

is the probability density function of the forward generation time (Britton and Tomba 2019, Champredon and Dushoff 2015, Wallinga and Lipsitch 2007). This representation separates the strength of transmission (*R*_0_) from the timing of infectiousness (captured by *f* (*a*)), enabling flexible investigation of both.

Modeling transmission in this form generalizes many epidemic models commonly used. For example, *S* usceptible-*I* nfectious-*R*ecovered (SIR) models presume constant transmission at rate *β* during an exponentially distributed infectious period with mean 1*/µ* (Anderson and May 1991). This can be captured by

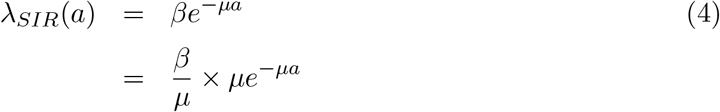

with *R*_0_ = *β/µ* and *f* (*a*) = *µe*^*−µa*^. Similarly, modifications of *S*usceptible-*E*xposed-*I*nfectious- *R*ecovered (SEIR) models have been widely applied to model SARS-CoV-2 transmission (Ferguson et al. 2020, Kissler et al. 2020, Morozova, Li and Crawford 2020). In such models, newly infected but not yet infectious persons enter an exposed state for an exponentially distributed length of time with mean 1*/µ*_1_, after which they become infectious for an exponentially distributed duration of mean 1*/µ*_2_ during which transmission again occurs at constant rate *β*. Letting *D*_1_ and *D*_2_ denote the duration of time after infection spent in the exposed and infectious states, early transmission in this model can be captured by

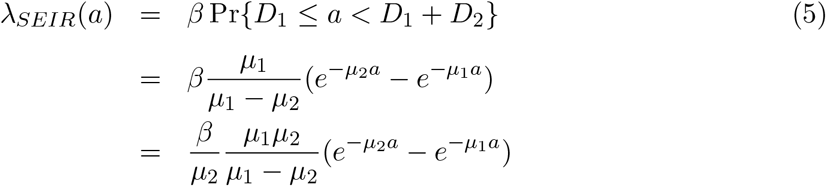

where *R*_0_ = *β/µ* _2_ and 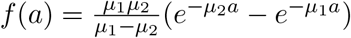.

Beyond SIR and SEIR models, epidemiologists have approximated generation time distributions directly from contact tracing data, and several such studies have been conducted using early SARS-CoV-2 outbreak data from China (see Park et al. 2020 for a summary). The generation times are often presumed to follow gamma distributions, as the latter provide a flexible statistical model for the time between the onset of symptoms for infector/infectee pairs within a transmission chain (the *serial interval*), and the distribution of serial intervals is taken as an estimate of the unobservable times between infections (which is what the generation time density *f* (*a*) is meant to represent).

## 3 Modeling the Impact of Testing and Isolation

Suppose that an infected student is isolated at age *T* days following infection, having been detected via repeat screening^1^. We model *T* as a random variable independent of the Poisson process of infections. Figure 1 shows the transmission rate *λ*(*a*) with the isolation age *T* represented by the vertical black line. The effect of isolation at *T* is to erase all infections that would have been transmitted beyond time *T*; this is illustrated as the shaded blue area in Figure 1.

**Figure 1:**
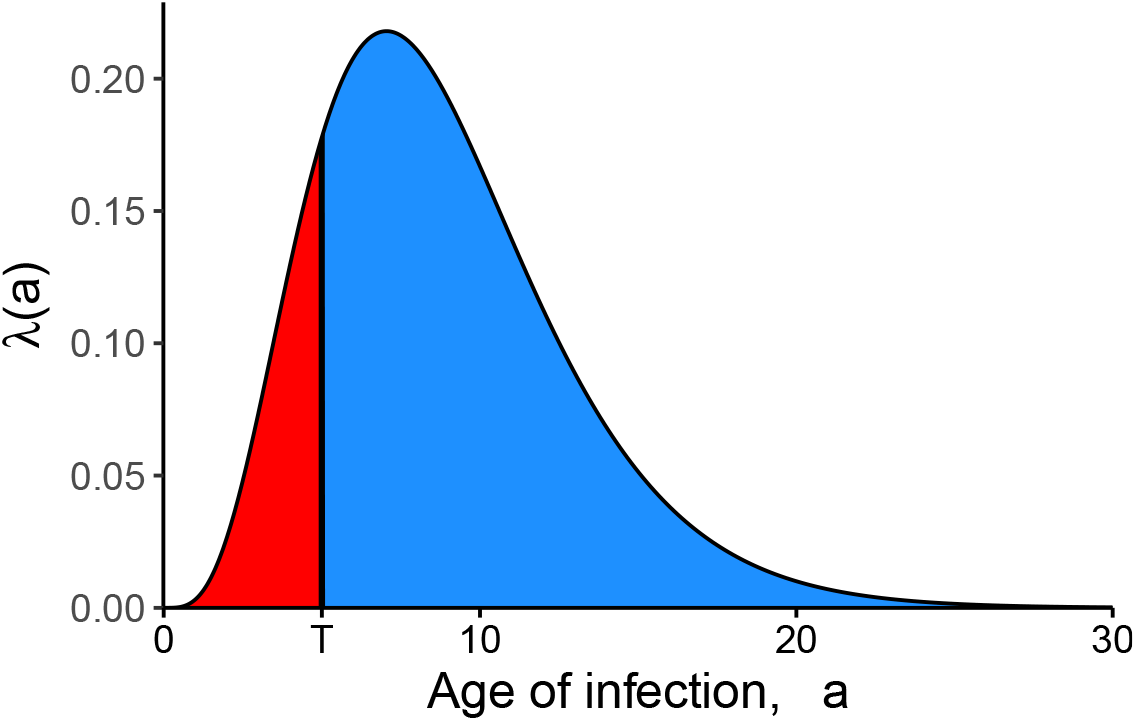
Impact of isolation. For a person isolated at a random time *T* after infection, the blue shaded area shows the expected number of further infections whose transmissions are prevented by the isolation, and the red area shows the expected number of further infections that escape isolation and are still transmitted.

The sooner an infectious person is isolated (the smaller the value of *T*), the greater the number of infections that can be prevented, and the fewer the number of transmissions that escape isolation. Following the Poisson model, conditional on *T*, the expected number of transmissions that occur before isolation is 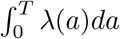. Thus, the expected number of infections that would escape isolation and still be transmitted, *R*_*T*_, is given by

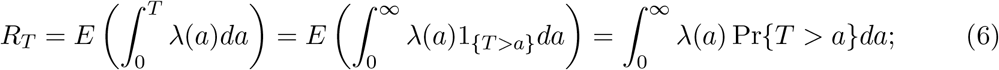

here 1_*B*_ denotes the indicator function taking the value 1 if the event *B* occurs and 0 otherwise. Defining *λ*_*T*_ (*a*) = *λ*(*a*)*P {T > a}* to be the *effective transmission rate* at age *a* taking account of the testing program, 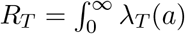*da* is the area under the curve *λ*_*T*_. Clearly *R*_*T*_ *≤ R*_0_ as Pr*{T > a} ≤* 1, with the extent of the reduction in transmission depending on the distribution of *T*, which in turn depends upon testing characteristics such as the timing of repeat tests, test sensitivity, and the lag time from testing to isolation.

### 3.1 Perfect Repeat Testing

As a contrast to the regularly-spaced testing that is the subject of most of this paper, consider a perfect test that on average is administered to each student once every *τ* days but whose timing is otherwise random with a constant hazard rate. This implies that *T* would follow an exponential distribution with mean *τ*, with

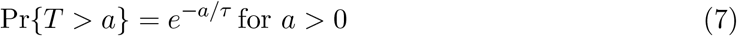

as a result. Alternatively, as a model of regularly scheduled testing, suppose that students are administered a perfect test literally once every *τ* days on a fixed schedule. For example, scheduled weekly testing would require each student to be tested once every seven days. One way to implement this would be for 1*/*7^*th*^ of the students to be tested each Sunday, a different 1*/*7^*th*^ each Monday, and so on such that each student has a specified day of the week (and time slot) for testing. For such a schedule in continuous time, *T* would follow a uniform distribution on (0, *τ*), and

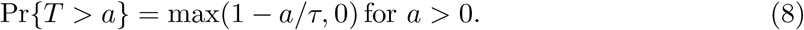

From equations (7) and (8), it is clear that scheduled screening would be more effective than random screening for all values of *τ* since *e*^*−a/τ*^ *>* 1 *− a/τ* for *a >* 0. With random screening, on average a newly infected person would not be detected and isolated until *τ* time units after infection, whereas with scheduled screening, the mean time from infection to isolation would be just *τ/*2, while *τ* would be the *maximum* time from infection until isolation (as opposed to the *mean* time with random screening). This distinction is important, as expanding SIR or SEIR models to include testing by applying a constant testing rate to the infected population amounts to random screening.

### 3.2 Imperfect Repeat Testing

Tests are not perfect, however. The sensitivity of a test is defined as the conditional probability of receiving a positive test result on an individual, given that the person tested is in fact infected. We denote the sensitivity of a screening test by *σ*. Random screening with a mean intertest period of *τ* also results in the time of detection being exponentially distributed, but now with mean *τ/σ*, inflating the time to detection by the factor 1*/σ*. Regular scheduled screening, with a deterministic separation of *τ* days between successive tests for a given individual, is more complicated. Let ⌊*x*⌋ denote the largest integer less than or equal to *x* (the *floor* function). Regular scheduled screening with sensitivity *σ* yields

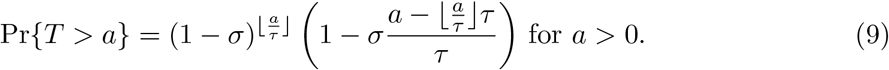

This follows because in each screening interval of duration *τ*, detection will occur with probability *σ*, which makes the number of testing intervals until the interval containing detection follow a geometric distribution with mean 1*/σ*. If detection occurs, the timing of detection within the interval will be uniformly distributed between 0 and *τ*. As a consequence, the expected time from infection to detection with scheduled imperfect screening once every *τ* days equals

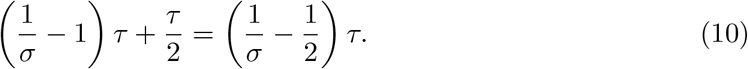

Note this time is shorter than the mean time to detection with imperfect random screening by *τ/*2 days, which is the same difference in mean detection times for scheduled versus random screening with perfect testing (*σ* = 1).

### 3.3 Scheduled Screening with Age-of-Infection-Dependent Sensitivity

Given both the shorter lags from infection to isolation and the ease of implementing scheduled versus random testing, we will narrow our focus to scheduled testing while increasing model realism. While we have included test sensitivity in our model, thus far we have presumed constant sensitivity that does not depend on the time since infection. This latter assumption is not realistic. For example, viral tests such as reverse transcriptase polymerase chain reaction (RT-PCR) cannot detect infections immediately after they occur, and indeed the ability of a test to detect the virus presumably behaves in a manner that is somewhat related to the ability of an infectious individual to transmit the virus (Kucirka et al. 2020). To model the dependence of sensitivity on time since infection, we denote the sensitivity of a test administered at an age of *a* time units after infection by *σ*(*a*), where *σ* is called the *sensitivity function* of the test. For simplicity we assume here that the results of tests taken at different times after infection are mutually independent given the sensitivity function.^2^

Determining the survivor function Pr*{T > a}* from the sensitivity function *σ*(*a*) is best approached by first deriving the probability density function *g*_*T*_ (*a*) for the isolation age *T*. In a scheduled repeat testing policy with screening interval *τ*, we want the probability that an individual who has been infected for *a* time units was not detected over the previous 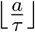 tests administered since becoming infected, but is tested and detected in the time slice (*a, a* + *da*). Owing to the independence of the infection, screening and detection processes, this probability is given by 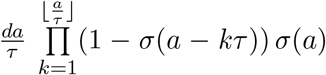, where an empty product equals 1 by definition. In other words, the probability density function for the time *T* from infection until isolation is given by

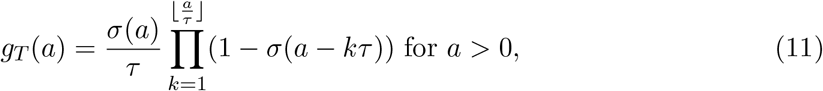

and the survivor function Pr*{T > a}* then follows from integration as

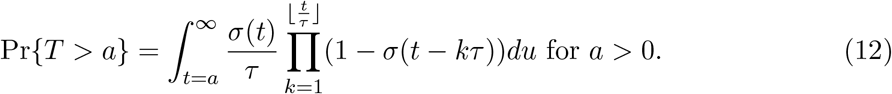

#### 3.3.1 Example: Step Function Sensitivity

One simple model of test sensitivity could be described by a *silent window* of duration *w* after infection during which it is not possible to detect the presence of infection, after which infection can be detected with constant sensitivity *σ* until time *r*, the *reach* of the test, beyond which the test becomes insensitive. In this case the test sensitivity would follow a step function over the time from infection, that is

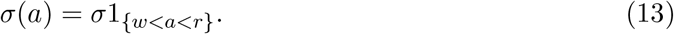

The survivor distribution Pr*{T > a}* is a function of *a ∧ r*, the minimum of *a* and *r*, and can be thought of as scheduled screening beginning at time *w* after infection (for no detection is possible within the window period). Due to the independence of the infection and testing processes, equation (9) still applies, but to the number of days since expiration of the window period rather than to the age of infection, so that the survivor distribution of the time from infection until detection is given by

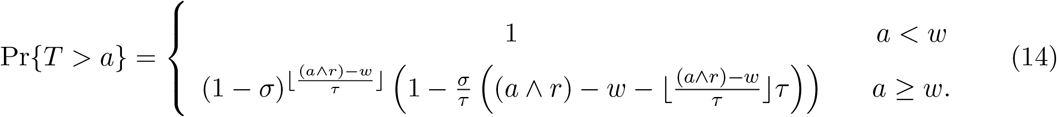

Accounting for the silent window *w* can substantially reduce the efficiency of repeat testing, as illustrated by the simple result in the case where the test reach *r* is infinite that the expected time from infection until detection increases by *w* days to 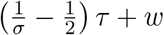.

#### 3.3.2 Example: Kucirka et al. (2020)

An estimated sensitivity function of reverse-transcriptase polymerase chain reaction (RT-PCR) tests for SARS-CoV-2 is provided by Kucirka et al. (2020), based on a Bayesian hierarchical model fit to data drawn from 7 previously published studies. Their sensitivity function is approximated by

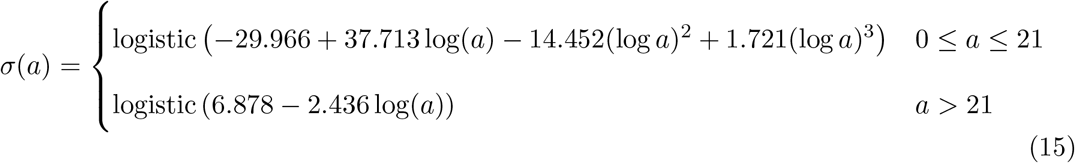

where logistic(*z*) = *e*^*z*^*/*(1 + *e*^*z*^) is the logistic function. This function fits precisely with values obtained by Kucirka et al. (2020) in the range 0 *≤ a ≤* 21, and then the cubic function of log(*a*) is extrapolated linearly on the logistic scale for *a >* 21.

### 3.4 Incorporating Delay From Testing To Isolation

Finally, tests take time to process, as does informing students of their test result and ensuring the start of isolation. We refer to this additional delay as the isolation lag, denoted by *ℓ*, and note that the impact of incorporating this lag is to shift any 0-lag survivor distribution for the time from infection to isolation by *ℓ* days to account for the additional delay. Define *T*_*ℓ*_ as the time from infection to isolation incorporating an isolation lag of *ℓ*, while *T*_0_ is the time from infection until isolation based on whatever screening interval *τ* or age-of-infection-dependent sensitivity *σ*(*a*) is being studied in the absence of isolation delay. Our final model for the survivor function for *T*_*ℓ*_, the time from infection until isolation accounting for the isolation lag, is given by

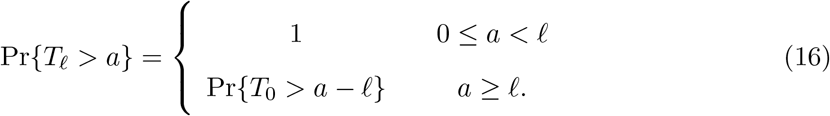

### 3.5 Illustrative examples

Figure 2 illustrates four examples of sensitivity functions:

**Figure 2:**
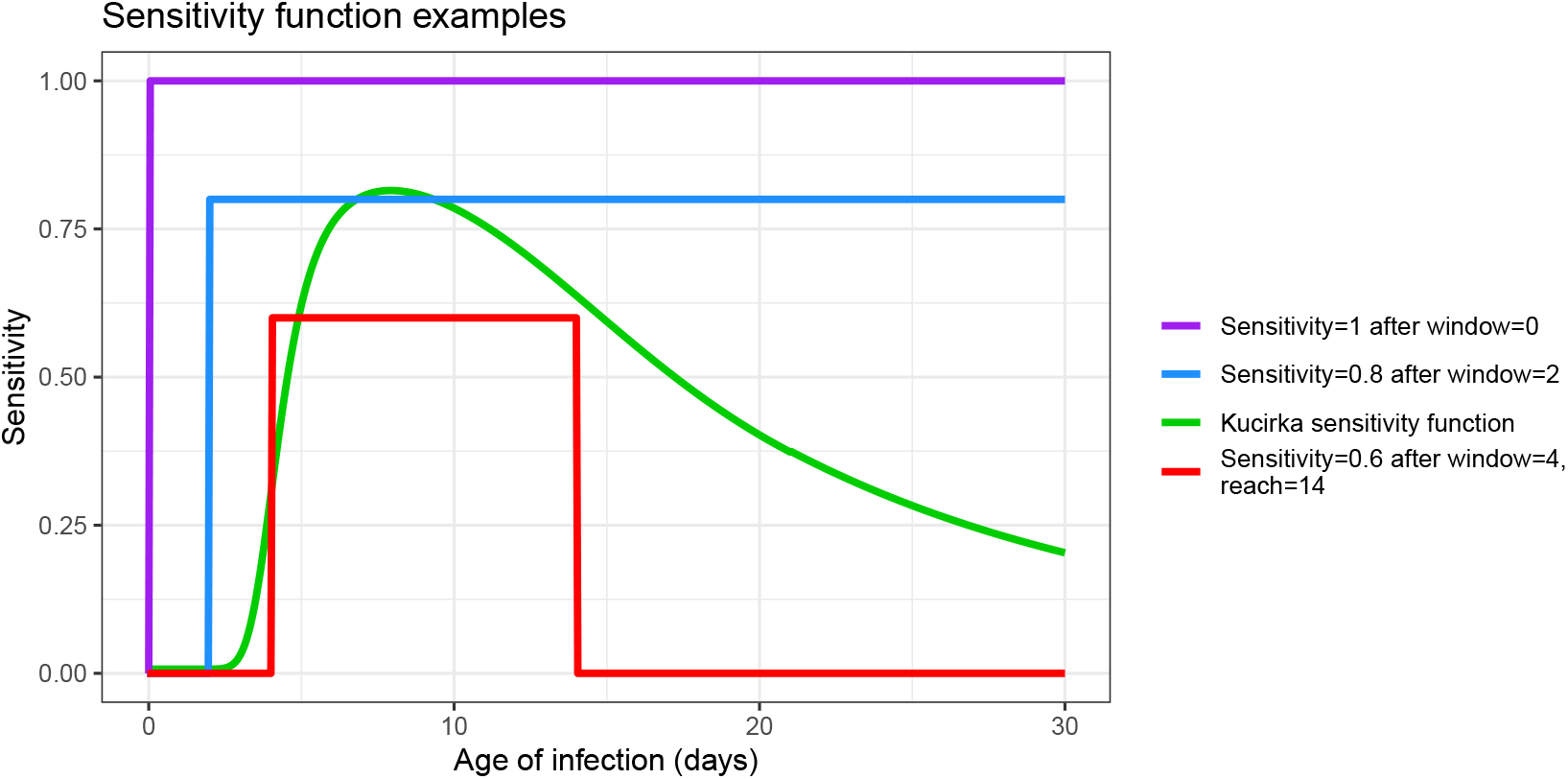
Examples of test sensitivity functions

1. perfect sensitivity (with no window of zero sensitivity)
2. a step function with sensitivity 0.8 (Hanson et al. 2020) after a window of 2 days with zero sensitivity,
3. the Kucirka et al. (2020) sensitivity function
4. a step function having a window of 4 days with zero sensitivity, followed by 10 days with sensitivity 0.6, after which the sensitivity returns to 0 (i.e. reach = 14 days).

Applying these four tests with regular weekly screening and 1 day of isolation delay in all cases, the corresponding survivor functions are shown in Figure 3. This figure shows how repeat testing is harmed by both imperfect testing (which forces multiple testing cycles to detect new infections), and an isolation lag (which shifts the survivor function one day to the right, increasing the time from detection to isolation).

**Figure 3:**
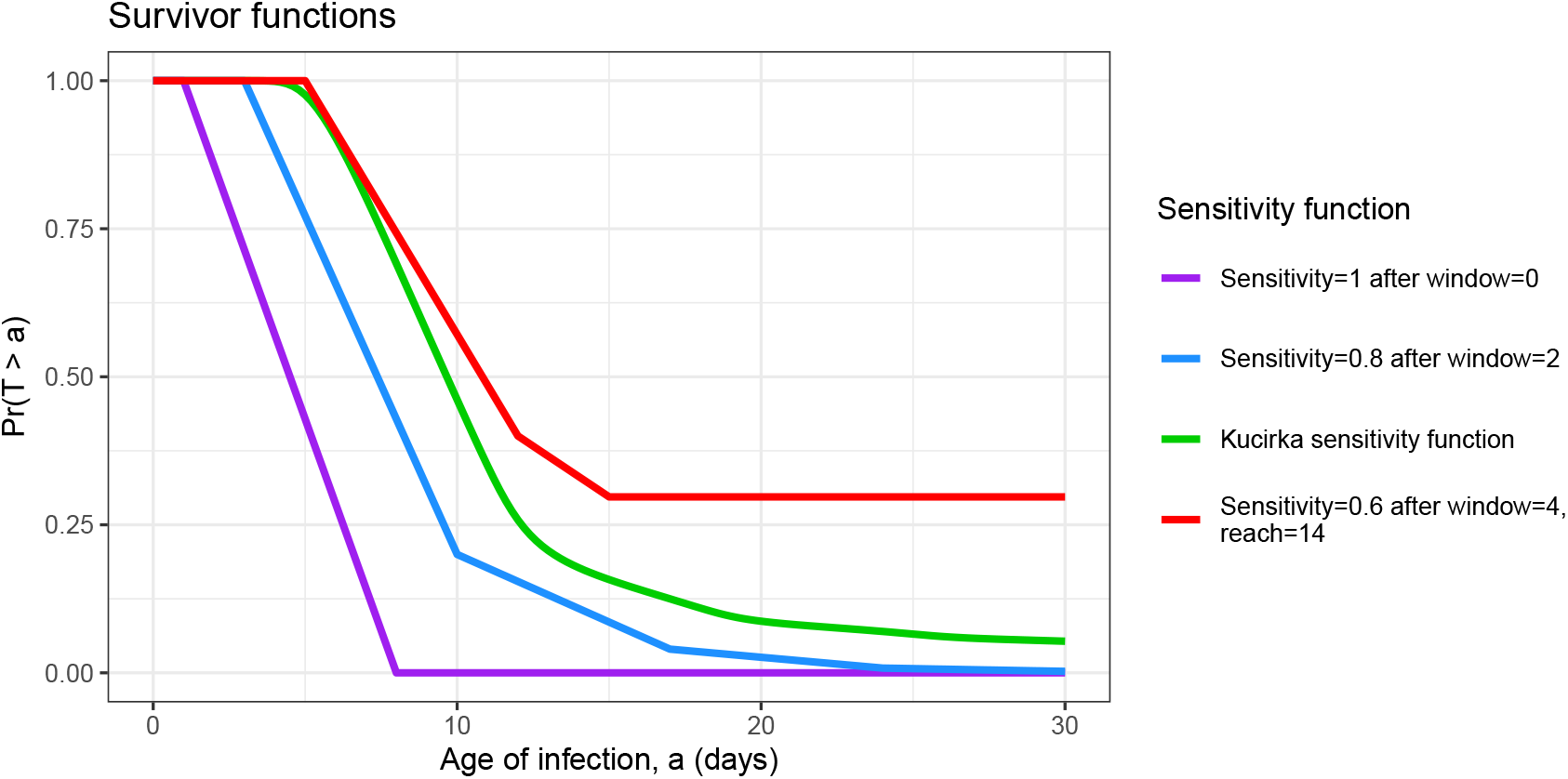
Probability that the time from infection to isolation exceeds *a*. Here the isolation delay in all four scenarios was taken to be 1 day.

Figure 4 plots the transmission function *λ*(*a*) corresponding to the forward generation time density implied by Li et al. (2020), which is a gamma distribution with a mean (standard deviation) of 8.86 (4.02) days, for an outbreak with *R*_0_ = 1.6. Also shown are the effective transmission curves found by multiplying by the four survivor functions of Figure 3.

**Figure 4:**
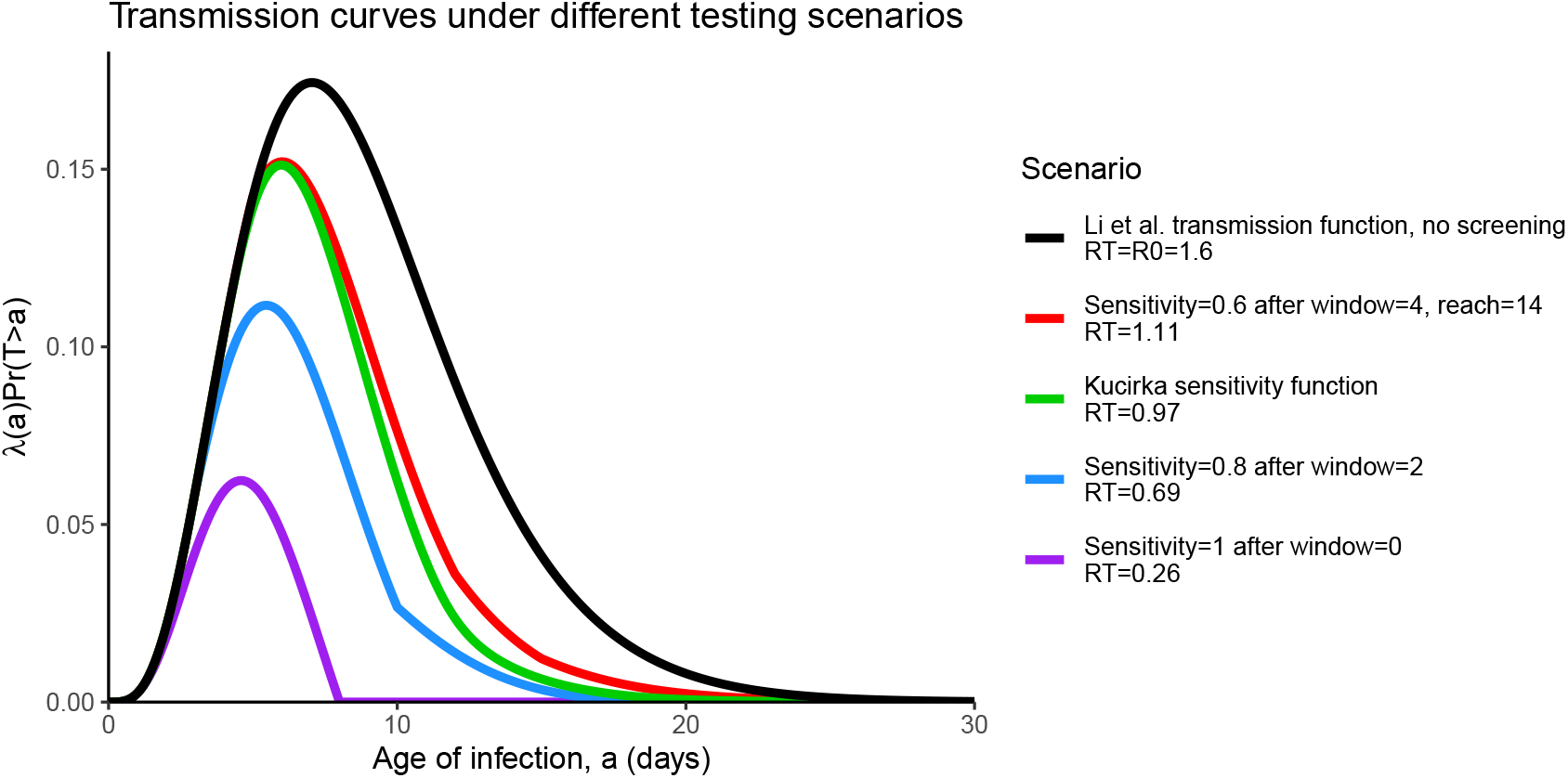
Transmission curves under different testing scenarios.

The effect of repeat testing on blocking transmission is considerable, but harmed by imperfect sensitivity and isolation delay. One way to quantify this effect is to compute *R*_*T*_ for each scenario; following equation (6), each *R*_*T*_ is the area under the corresponding effective transmission curve. Starting with *R*_0_ = 1.6 in the absence of any screening, testing students each week with perfect sensitivity would result in *R*_*T*_ = 0.26. Replacing perfect screening by a window of 2 days of zero sensitivity followed by sensitivity 0.8 increases *R*_*T*_ to 0.69. Replacing the sensitivity function by that of Kucirka et al. (2020) increases *R*_*T*_ to 0.97. The fourth example sensitivity function with lower sensitivity, longer window of zero sensitivity, and finite test reach further increases *R*_*T*_ to 1.11; the effect of the test reach is relatively minor though since it changes the survivor function at times *a* large enough so that *λ*(*a*) is relatively small. This shows that weekly screening helps reduce transmission, but is harmed by imperfect test sensitivity in ways that depend on the shape of the sensitivity function.

## 4 Dynamic Transmission Model

To expand this framework to a dynamic transmission model, we follow Kaplan (2020b) with slightly different notation while incorporating the effect of repeat screening and define:

*s*(*t*) *≡* fraction of the population that is susceptible to infection at calendar time *t*;

*π*(*t*) = incidence of infection at time *t*;

*λ*(*a*) *≡* the transmission intensity as a function of age of infection introduced earlier;

*T* = the isolation age induced by repeat testing as discussed previously with distribution characterized by the survivor function Pr*{T > a}*

Thinking of time 0 as the start of the term when students arrive to campus, given initial conditions, the dynamic screening model can be written as:

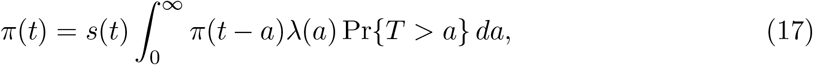

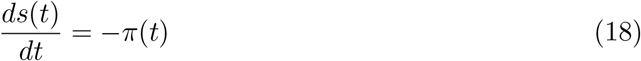

for *t >* 0. Equation (17) sets SARS-CoV-2 incidence proportional to the fraction of the population that is susceptible and the infected population-weighted age-of-infection adjusted transmission intensity thinned by the effect of repeat testing and equation (18) depletes susceptibles with the incidence of infection. Note that the distribution of infectiousness over time from infection is implicitly accounted for in the definition of *λ*(*a*), thus there is no need for explicit removal of infectious persons from the population as they simply cease transmitting in accord with *λ*(*a*).

To specify initial conditions for the model we suppose each student is tested in an initial screening. Letting *π*_0_ denote the fraction of students who were infectious at time 0 but not detected by the initial screening, we assume that their ages of infection at time 0 are uniformly distributed over an interval [0, *A*], so that *π*(*t*) = *π*_0_*/A* for *t ∈* [*−A*, 0]. We choose *A* large enough so that it is a good approximation to consider students at time 0 with infections of age greater than *A* as no longer infectious. With this assumption there is no need to keep track of when infections of age greater than *A* at time 0 occurred, but rather it is enough to note their total number as reflected in the initial susceptibility *s*(0). Thus initial conditions are given by *s*(0) and *π*_0_.

### 4.1 Incorporating Imported Infections

Thus far the model has only considered the detection of internally generated infections due to a closed outbreak beginning with the initial conditions *s*(0) and *π*_0_. However, due to off-campus wanderings as well as visitors to campus, one can expect on-campus residents to be infected by external exposures. A screening policy must contain transmission generated by such imported infections in addition to internal transmission among college residents. Let *v*(*t*) denote the exposure rate of imported infections at time *t* per campus resident. The rate such exposures lead to actual infections presuming on-campus residents are exposed at random then equals *v*(*t*)*s*(*t*).

For example, if there are *n* on-campus residents, and on average one such resident has one imported exposure sufficient to transmit infection weekly (either by direct off-campus exposure or as the result of exposure to an infected visitor on-campus), then *v*(*t*) = 1*/*(7*n*) per day. If instead a single sufficient imported exposure happens on a daily basis, then *v*(*t*) = 1*/n* per day. We modify our model by including transmission from imported infections in the on-campus incidence rate, and thus modify equation (17) to

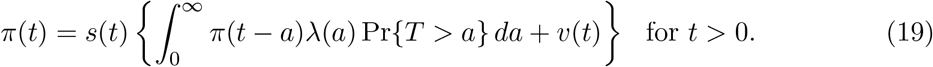

Note the dual role played by *v*(*t*): imported infections will contribute to on-campus incidence the same way infections acquired on-campus contribute over time, marking *transmission from* imported infections. But the persons who *acquired* these imported infections immediately deplete the on-campus susceptible population at their time of infection. Both effects are accounted for in equation (19); *v*(*t*)*s*(*t*) is the instantaneous contribution to incidence by imported exposures at time *t*, and via equation (18) immediately contribute to the depletion of susceptibles.

### 4.2 Performance Measures: Cumulative Incidence, Isolation, and Undetected Infections

The cumulative incidence *c*(*h*) of infections that occur over some planning horizon *h* is given by

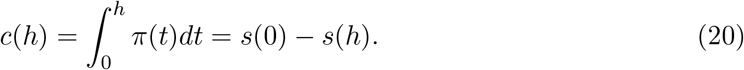

Minimizing transmission is the most important goal of a repeat testing program, but it is not the only one. Administrators will also need to have an estimate of the number of students that screening will detect and isolate. Until this point in our discussion, we have focused on detecting actual infections, that is, true positives, but testing also produces false positive errors (Hanson et al. 2020) that will land additional students in isolation. We now consider both true and false positives in determining the number of students who would require isolation over the planning horizon.

Let *δ*_*T P*_ (*t*) denote the true positive isolation rate, that is, the rate at which infected students are isolated accounting for scheduled screening frequency, test sensitivity, window and reporting lag at time *t* from the start of the planning horizon. This isolation rate is given by

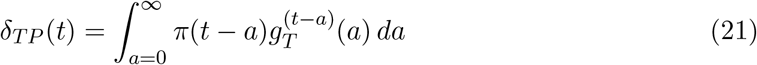

where 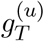 denotes the probability density function for the isolation age *T* for an infection that occurred at time *u*. This carries some dependence on *u* because in our model we assume that regular screening begins at time 0 so that an individual infected at time *u <* 0 is not tested for the first |*u*| units of time following infection. The density 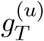 may be obtained by the general equation (11) applied to a test sensitivity function that has been modified by multiplying it by the indicator function 1*{a>*|*u*|*}*, since we can think of not being tested for the first |*u*| time units after infection as equivalent to using a test that has sensitivity 0 for the first |*u*| time units. In our calculations we approximate *δ*_*T P*_ (*t*) by replacing the infinite upper limit of integration in (21) by *A*.

To model the false positive isolation rate *δ*_*F P*_ (*t*), let *ϕ* denote the false positive rate of the test (which equals one minus the *specificity*). To become a false positive isolated at time *t* a person needs to be susceptible, tested at time *t − ℓ*, and receive a false positive error on the test, which suggests

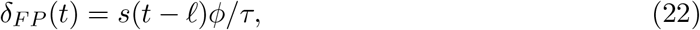

where *ℓ* is the isolation lag and *τ* is the spacing of the regular tests.

Students testing positive thus enter isolation at time *t* with total rate *δ*(*t*) = *δ*_*T P*_ (*t*) + *δ*_*F P*_ (*t*), and remain isolated for duration Δ. The fraction of the population in isolation at time *t, ι*(*t*), when the duration of isolation is equal to Δ (typically 14 days) thus equals

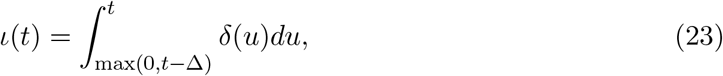

with corresponding formulas for true positive and false positive isolations, *ι*_*T P*_ and *ι*_*F P*_, in terms of the functions *δ*_*T P*_ and *δ*_*F P*_. We assume that false positive detections are not susceptible while in isolation but then they return to the susceptible pool and to regular testing once they leave isolation.

Finally, integrating equation (21) yields the total fraction of the population that was infected and detected over the course of the outbreak. Comparing this result to the cumulative incidence in the population yields the fraction of the population that was infected but not detected.

## 5 Scenario Analyses and Outbreak Control

The models described throughout this paper have been implemented in a web app available at https://jtwchang.shinyapps.io/testing/. The app allows the user to select values from a wide range of model input parameters as illustrated in Figure 5. The app also allows users to address the timing of transmission as implied by the forward generation time density *f* (*a*). We consider two different models for *f* (*a*). The first is the gamma distribution with mean 8.87 days and standard deviation 4.02 days, drawn from Li et al.’s (2020) study of early transmission dynamics in Wuhan referred to earlier, which is widely cited as the first detailed analysis of early SARS-CoV-2 transmission. The second is also a gamma distribution but with mean (standard deviation) equal to 8.50 (6.07) days; this is based on parameter point estimates in the Bayesian meta-analysis conducted by Park et al. (2020). These two forward generation time densities are displayed in Figure 6. Comparing these distributions, we see that while the Park et al. and Li et al. densities have similar means of 8.9 and 8.5 days, the standard deviation is smaller for the Li density, causing generation times to cluster closer to the mean which implies delayed transmission. The Park et al. density rises more steeply and peaks earlier, presenting an early transmission challenge.

**Figure 5:**
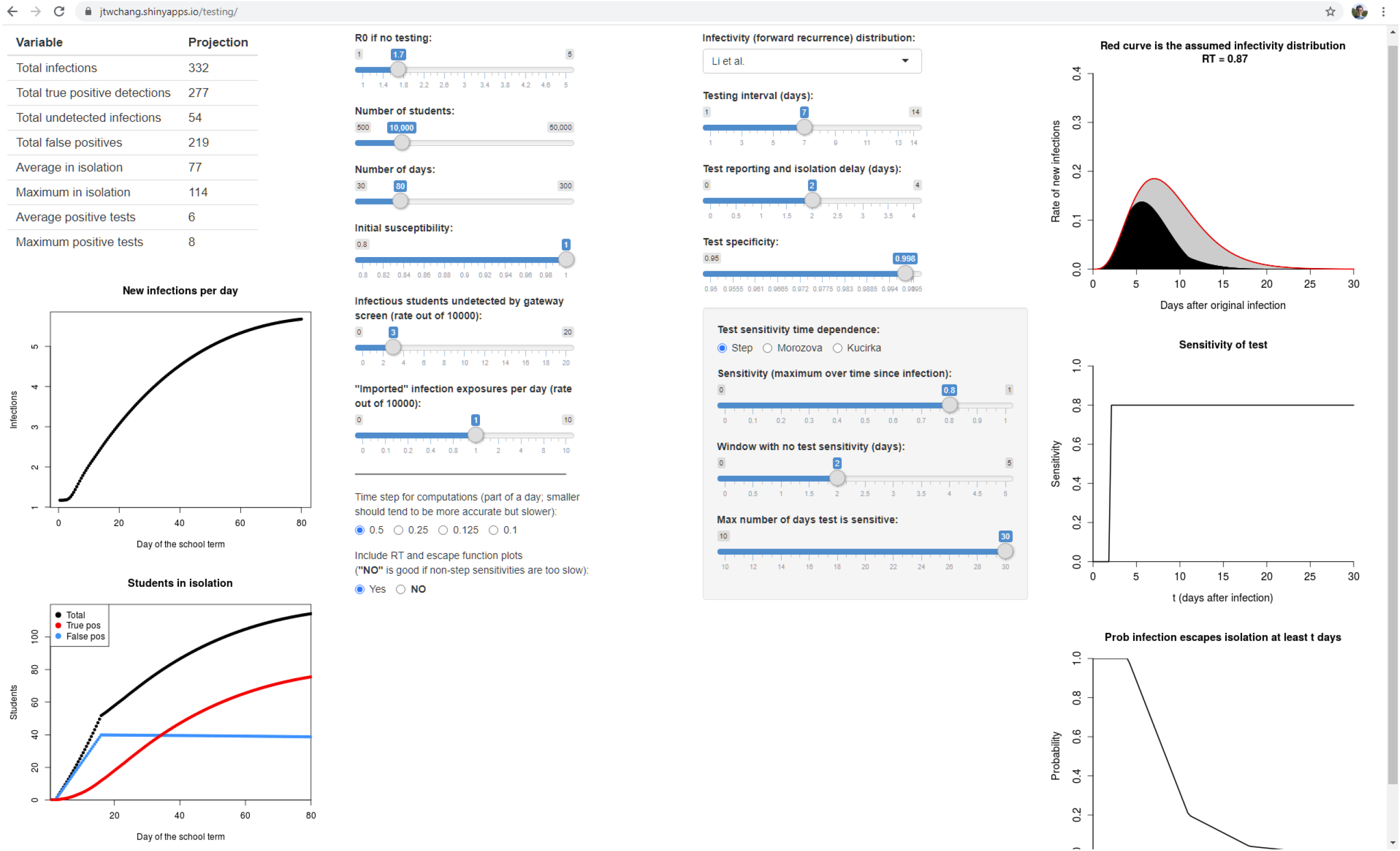
A web app available at https://jtwchang.shinyapps.io/testing/ that implements the model and facilitates exploring a variety of scenarios.

**Figure 6:**
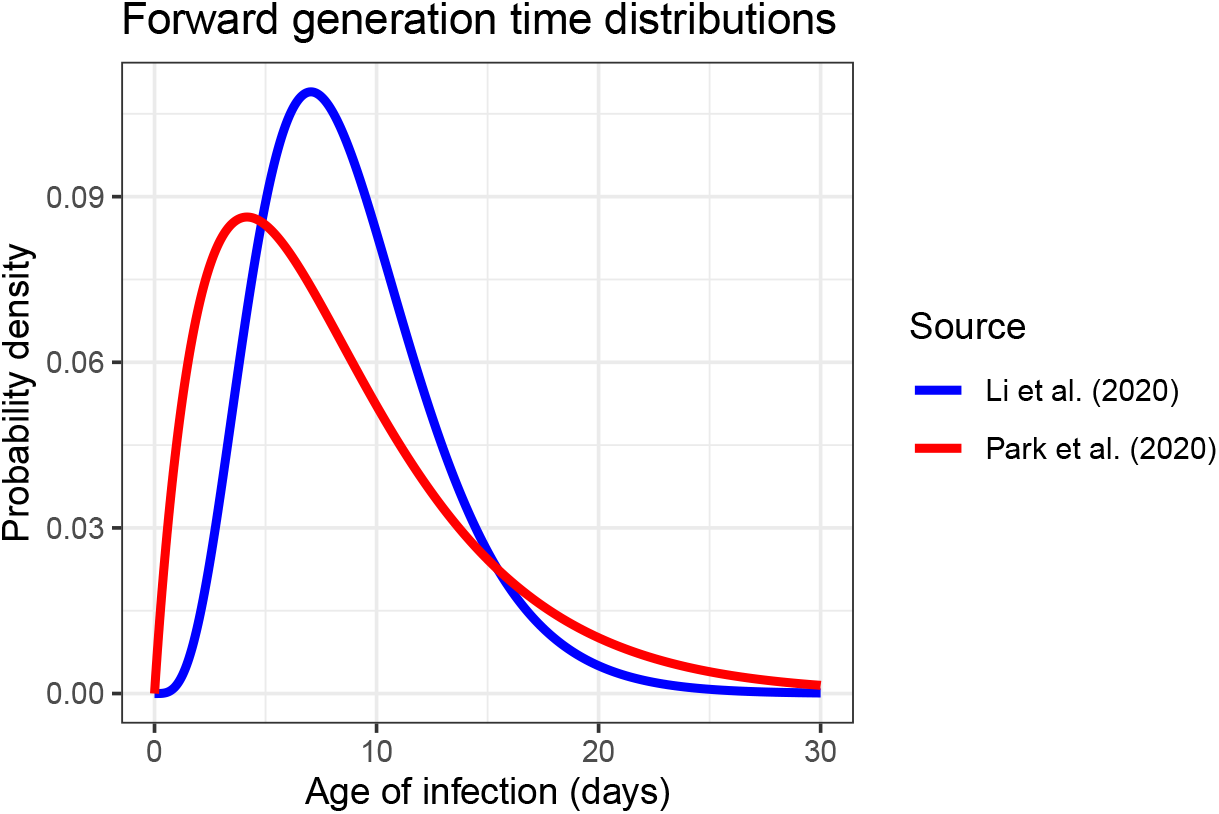
Two estimated generation time distributions found in published studies. We refer to these distributions as featuring relatively *early transmission* (Park et al. 2020) and *late transmission* (Li et al. 2020).

We illustrate the model with four testing scenarios over an 80 day period simulating an abbreviated fall term in a population of 10,000 students with reproductive numbers of 1.0, 1.5, 2.0, and 2.5 using the Li et al. (2020) forward generation time distribution. We assume that testing takes place every three days, set *v*(*t*) = 1 imported exposure per day, test specificity equals 99.8% (Hanson et al. 2020), and test sensitivity follows the trajectory estimated by Kucirka et al. (2020) discussed earlier. The outbreaks begin with three initially infectious students at the start of school (everyone else in the population is susceptible), and a 24 hour delay from testing to student isolation for students testing positive. Figure 7 plots the cumulative number of infections in these four scenarios.

**Figure 7:**
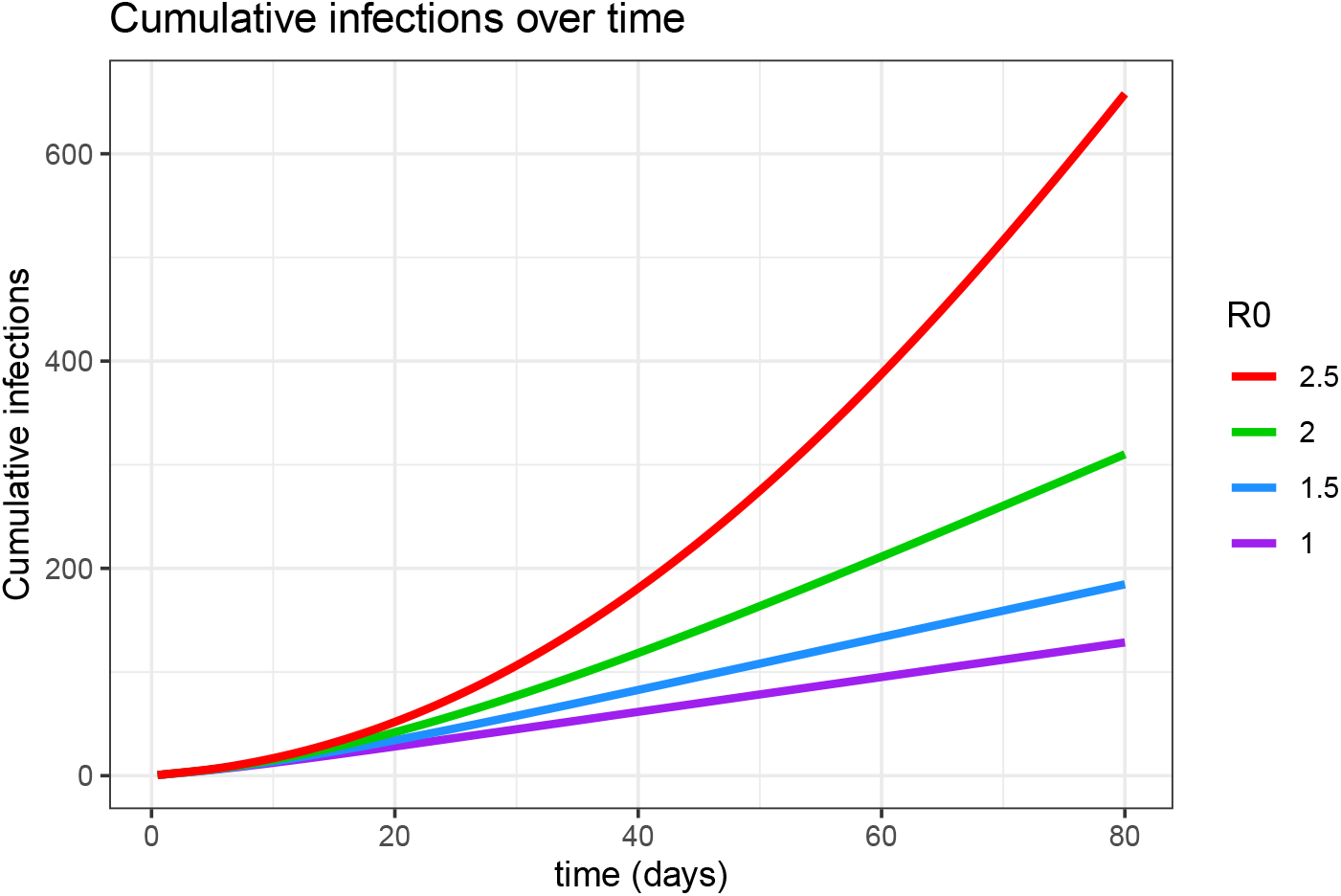
Cumulative infections over time in scenarios with testing every 3 days and fixed infectivity function, sensitivity function, and delays, as *R*_0_ varies.

Cumulative infections increase from 131, to 187, to 312, and then 658 as *R*_0_ increases from 1.0 to 2.5 in increments of 0.5, while the time averaged number of students isolated equals 102, 109, 124, and 162 in these same scenarios. There are about 90 false positives in isolation on average in all four scenarios. The nonlinear effect of *R*_0_ on these otherwise equivalent scenarios is notable. University preparations in the realms of social distancing and infection control are meant to lower *R*_0_, and if students comply with such directives, the likelihood of achieving a favorable epidemic outcome should increase. However, some are skeptical that students will comply with such directives (Steinberg 2020), which could lead to higher reproductive numbers (and imported exposures) and worse epidemic outcomes, perhaps comparable to cruise ship transmission (Zhang et al. 2020).

This model is flexible in allowing users to simulate many different testing scenarios, but it is perhaps most useful in identifying the limits of outbreak control for alternative repeat-testing policies. The proposed approach is to first identify an acceptable control level of infection within a defined time periord, such as 5% of the student population over the course of a semester. Such a control level could reflect the maximum number of infections university health systems can handle considering realistic testing (both collection and laboratory resources) and isolation capacity (residential space, human resources for monitoring, counseling and compliance). The control level could also reflect university concern with secondary transmission from students to vulnerable persons such as certain faculty, workers, or the residents of the surrounding community in which the university is embedded. The control level could even follow from a mortality goal such as ensuring the probability of zero COVID-19 fatalities is at least 99%.

For a given repeat testing interval, one can use the app to determine the most challenging parameter values for which total infections remain within the previously stated control level. Repeating this for different testing frequencies thus helps determine the limits of control for each policy. While identifying appropriate control limits is the responsibility of university leadership as opposed to analysts, having the ability to show officials the limits of different control strategies enables senior decision makers to trade off infection outcomes against other important considerations including testing costs as well as intangibles such as the importance and value of residential education in the midst of a pandemic.

We illustrate by again considering a scenario where 10,000 students will be repeatedly tested over 80 days. We maintain the assumptions that there is one imported exposure per day, test specificity equals 99.8%, there are three initially infectious students, and a 24 hour delay from testing to student isolation. There are four transmission and detection scenarios considered, corresponding to using the late-transmission Li et al. (2020) or early-transmission Park et al. (2020) forward generation time density with either the Kucirka et al. (2020) or step-function sensitivity, where the step-function presumes a two day non-detection window followed by constant 80% sensitivity (Hanson et al. 2020). For weekly screening and testing every three days, we determine the largest value of *R*_0_ (in increments of 0.05) such that total infections are held beneath 500 (or 5% of the population tested), and report total infections, average and maximum daily numbers of students isolated, and average daily positive tests.

Table 1 reports the maximal values of *R*_0_ for weekly screening that can keep infections below 5% of the population. The most pessimistic scenario -- early transmission and Kucirka et al. (2020) sensitivity -- requires that *R*_0_ falls at 1.4 or below. The most optimistic scenario -- late transmission and the presumed step-function sensitivity -- keeps infections below 5% providing *R*_0_ falls below 2.25. The two intermediate scenarios contain infections below 5% of the population providing *R*_0_ is at most 1.6-1.8. While all four of these scenarios result in comparable numbers of infections and daily positive tests, note that both scenarios employing the step-function sensitivty on average isolate more students than the remaining scenarios. This is because of the high 80% test sensitivity that applies once the two-day non-detection window expires in the step-function scenarios. A greater number of infected students are detected as a consequence, leading to the larger number of students in isolation. CDC (2020) recommends considering *R*_0_ to fall in the range from 2 to 3 in modeling studies, with 2.5 serving as their recommended base case value. Our analysis suggests that weekly testing could not contain infections below 5% for CDC’s base case reproductive number. However, the CDC recommendations are not specifically for residential college outbreaks, where one would hope that social distancing and infection control protocols would result in milder outbreaks with lower values of *R*_0_. On the other hand, conservative planning principles would suggest that hope is not enough, especially given recent evidence regarding outbreaks already occurring at residential colleges (Ellis 2020; Fields 2020). The wide range of results reported in Table 1 suggests that while weekly screening could contain an otherwise large-scale outbreak under favorable conditions of late transmission and (relatively) early detection with 80% test sensitivity, overall weekly screening is not sufficiently robust to reliably contain outbreaks in the residential college setting.

**Table 1:** Weekly Screening Results for an 80-day term for various scenarios described in the text.

| Scenario<br>($f(a)$, $\sigma(a)$) | Maximal $R_0$ | Infections | Average<br>Isolated | Maximum<br>Isolated | Positive<br>Tests/day |
| --- | --- | --- | --- | --- | --- |
| Li et al.<br>Kucirka | 1.6 | 472 | 87 | 152 | 7 |
| Li et al.<br>step-function | 2.25 | 465 | 93 | 155 | 8 |
| Park et al.<br>Kucirka | 1.4 | 447 | 87 | 139 | 7 |
| Park et al.<br>step-function | 1.8 | 456 | 99 | 156 | 8 |

Table 2 reports results for testing once every three days. The worst case scenario combining early transmission with Kucirka et al. (2020) sensitivity can now contain outbreaks for any reproductive number lower than 1.75, while the optimistic scenario combining late transmission with step-function sensitivity could contain outbreaks with *R*_0_ as large as 4.8. The intermediate scenarios can keep infections below 5% of the population for reproductive numbers as large as 2.3-2.65. Of course, compared to weekly screening, the number of students isolated increases greatly due to the inevitable increase in false positives associated with more frequent testing. Testing students every three days is thus more robust than weekly screening in that the range of reproductive numbers for which infections can be kept below 5% is larger for all scenarios. Such improved performance comes at the expense of isolating many more students over the semester, in addition to the cost of the increased number of tests required.

**Table 2:**
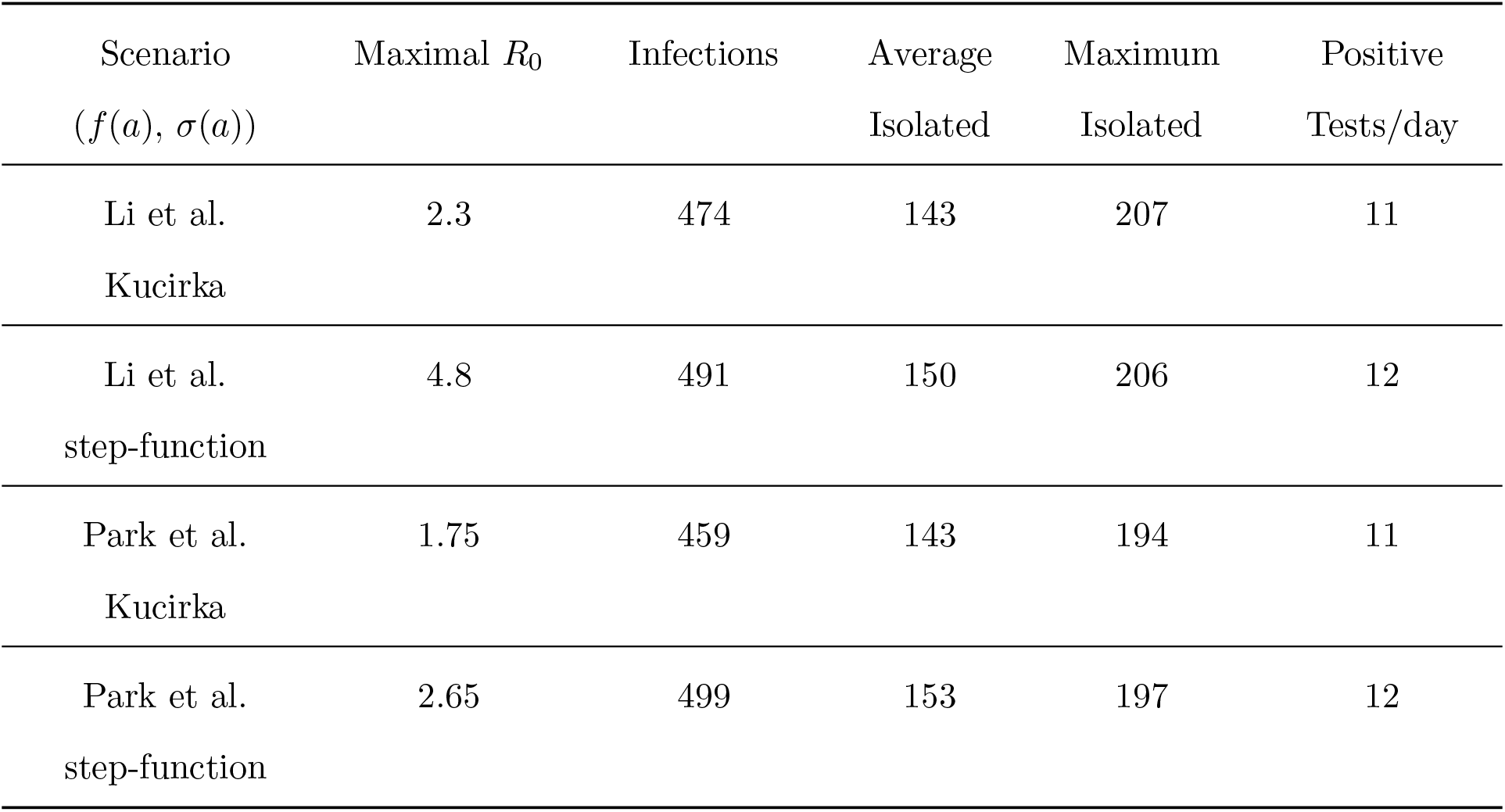
Testing Every Three Days. Results when testing every 3 days replaces the weekly testing in the scenarios of Table 1.

| Scenario<br>($f(a)$, $\sigma(a)$) | Maximal $R_0$ | Infections | Average<br>Isolated | Maximum<br>Isolated | Positive<br>Tests/day |
| --- | --- | --- | --- | --- | --- |
| Li et al.<br>Kucirka | 2.3 | 474 | 143 | 207 | 11 |
| Li et al.<br>step-function | 4.8 | 491 | 150 | 206 | 12 |
| Park et al.<br>Kucirka | 1.75 | 459 | 143 | 194 | 11 |
| Park et al.<br>step-function | 2.65 | 499 | 153 | 197 | 12 |

These examples illustrate how, other things being equal, more frequent screening enables adequate infection control to be achieved over wider ranges of values of *R*_0_. The examples also indicate that it is more difficult to contain scenarios where more transmission occurs earlier after infection (as with the Park et al. (2020) generation time distribution) rather than later (as with Li et al. (2020)). Another factor that can make infection control more difficult is the rate of imported exposures. Both Tables 1 and 2 presumed a single daily imported exposure over the modeled outbreaks; increasing this rate can make matters much worse. For example, for the Li et al. (2020) / Kucirka et al. (2020) scenario when testing every 3 days shown in the first row of Table 2, if the rate of imported exposures were to increase from 1 to 2 out of 10000 students, the maximal *R*_0_ for which infections could be kept below 500 would decrease from 2.3 to 1.8. Another factor of key importance is the delay from testing until isolation for those receiving a positive test result. For example, again in the example of the first row of Table 2, if the delay from testing to isolation increased from 24 to 48 hours, the maximal *R*_0_ at which infections could be kept below 500 would drop from 2.3 to 1.9, and adding one more day to increase the delay to 72 hours would further reduce the maximal controllable *R*_0_ to 1.65. This shows that two additional days of post-test delay would render testing once every three days no more effective than weekly testing with one day of delay.

## 6 Discussion

With much of the world only now emerging from COVID-19 lockdowns, educational institutions are struggling with a fundamental question: absent a vaccine against SARS-CoV-2 or an effective treatment for COVID-19, is it safe to bring residential students back to campus? Presuming infections can enter the student population, and recognizing that many if not most such infections will be asymptomatic, the ability to detect and isolate infections as they occur is crucial to prevent large outbreaks among students on campus and ignited by students off campus. Testing itself is not a panacea; it is the isolation of infectious students that prevents transmission, and should isolation not follow the detection of infected students, repeat screening would be relegated to producing descriptive outbreak statistics rather than actively stopping outbreaks from happening. This article has shown how repeat testing interrupts transmission via the isolation of infectious students, and analyzed numerous transmission scenarios.

We hope that this model and its web-based implementation are useful in helping college officials assess and anticipate quantitative influences of key factors that affect the performance of repeat screening programs. In particular, with substantial uncertainty surrounding multiple model inputs, it is prudent to explore a range of plausible scenarios, and it quickly becomes clear that plausible scenarios exhibit a wide range of outcomes from well controlled to badly out of control. While uncertainty and imprecise knowledge of inputs to our model preclude precise projections of future results, we can draw insights from the modeling that can help inform planning and implementation. For example, delay from testing until isolation emerges as a key target for control as we see how much each day of delay is expected to degrade the infection control benefits that high-frequency repeat testing can bring.

This analysis suggests that administrators must proceed cautiously and with open eyes when designing residential college screening programs, for while repeat testing for SARS-CoV- 2 infection can be a powerful tool for preventing infections and preserving public health in the residential college setting, it is not guaranteed to succeed. Even if students are tested once every three days, there are plausible transmission scenarios where the model projection has 5% or more of a student population becoming infected over the course of an abbreviated 80 day semester. Unlike engineering systems that are built conservatively to withstand multiple failures, the repeat testing system is necessarily fragile in that to succeed, all of the system components must work. Students must comply with infection control, social distancing, test scheduling and (if testing positive) isolation requirements for the repeat testing system to work effectively. The tests themselves must perform at or above expectation in terms of their ability to detect infected students. Isolation delay, including laboratory turnaround time, must be minimized as extra delay markedly degrades the ability of repeated testing to control outbreaks. While many of the factors involved are beyond control, college administrators should be able to implement systems that minimize isolation delay, both by contracting with testing laboratories to guarantee acceptable test turnaround times and by putting in place efficient communications and support mechanisms so that students who do test positive can be isolated as quickly as possible. Colleges can also effectively inform students what behaviors will be expected of them on campus while also clearly communicating the consequences of failing to comply with the adopted behavioral code. Finally, if a repeat testing and isolation program begins to lose control and infections are detected at higher rates than anticipated, colleges can shut down and confine students to quarters while ensuring that all those in need of medical attention receive it. The whole point of repeat screening is to avoid such an outcome, but nonetheless university administrators must be ready to close their residential colleges should repeat testing fail to contain the spread of SARS-CoV-2 on campus.

## Data Availability

Readers may explore the model and data employed via the app in the data availability link.

https://jtwchang.shinyapps.io/testing/

## Footnotes

1 Isolation would typically last only two weeks, but incorporating this would modify our results only slightly while complicating the analysis; see Kaplan (2020a) for details.

2 This independence assumption could be generalized, and in fact the only probabilities the current model would require are of the form Pr*{R*_*t*_ = *R*_*t*+*τ*_ = … = *R*_*t*+*kτ*_ = 0*}* where *R*_*t*_ is the result (1 for positive and 0 for negative) of a test at time *t* after infection for a given person.

